# Clinical Variables Correlate with Serum Neutralizing Antibody Titers after COVID-19 mRNA Vaccination in an Adult, US-based Population

**DOI:** 10.1101/2022.04.03.22273355

**Authors:** Min Zhao, Rebecca Slotkin, Amar H. Sheth, Lauren Pischel, Tassos C. Kyriakides, Brinda Emu, Cynthia McNamara, Qiaosu Shi, Jaden Delgobbo, Jin Xu, Elizabeth Marhoffer, Aleagia Mercer-Falkoff, Jürgen Holleck, David Ardito, Richard E. Sutton, Shaili Gupta

## Abstract

**Background:** We studied whether comorbid conditions impact strength and duration of immune responses after SARS-CoV-2 mRNA vaccination in a US-based, adult population.

**Methods:** Sera (pre-and-post-BNT162b2 vaccination) were tested serially up to 12 months after two doses of vaccine for SARS-CoV-2-anti-Spike neutralizing capacity by pseudotyping assay in 124 individuals; neutralizing titers were correlated to clinical variables with multivariate regression. Post-booster (third dose) effect was measured at 1 and 3 months in 72 and 88 subjects respectively.

**Results:** After completion of primary vaccine series, neutralizing antibody IC50 values were high at one month (14-fold increase from pre-vaccination), declined at six months (3.3-fold increase), and increased at one month post-booster (41.5-fold increase). Three months post-booster, IC50 decreased in COVID-naïve individuals (18-fold increase) and increased in prior COVID-19+ individuals (132-fold increase). Age >65 years (β=-0.94, p=0.001) and malignancy (β=-0.88, p=0.002) reduced strength of response at 1 month. Both strength and durability of response at 6 months, respectively, were negatively impacted by end-stage renal disease [(β=-1.10, p=0.004); (β=-0.66, p=0.014)], diabetes mellitus [(β=-0.57, p=0.032); (β=-0.44, p=0.028)], and systemic steroid use [(β=-0.066, p=0.032); (β=-0.55, p=0.037)]. Post-booster IC50 was robust against WA-1 and B.1.617.2, but the immune response decreased with malignancy (β =-0.68, p=0.03) and increased with prior COVID-19 (p-value < 0.0001).

**Conclusion:** Multiple clinical factors impact the strength and duration of neutralization response post-primary series vaccination, but not the post-booster dose strength. Prior COVID-19 infection enhances the booster-dose response except in individuals with malignancy, suggesting a need for clinically guiding vaccine dosing regimens.

**Summary:** Multiple clinical factors impact the strength and duration of neutralization response post-primary series vaccination. All subjects, irrespective of prior COVID infection, benefited from a third dose. Malignancy decreased response following third dose, suggesting the importance of clinically guided vaccine regimens.

## 1. Introduction

Severe Acute Respiratory Syndrome Coronavirus-2 (SARS-CoV-2) virus has infected more than 508 million people worldwide as of April, 2022, and has caused over six million deaths.^1^ Three vaccines have been approved by the United States Food and Drug Administration, based on safety and efficacy.^2^ The mRNA vaccines encode a codon-optimized version of spike glycoprotein (S) of SARS-CoV-2, which is the viral protein that elicits neutralizing antibodies that block viral entry into cells and subsequent replication. ^3–5^ Neutralizing antibodies to the spike protein of SARS-CoV-2 correlate with immunity against the virus after vaccination, reducing rates of COVID-related infection, hospitalization, and death.^6,7^ Neutralizing antibody titers have been shown to decline over a period of 6 months after completion of the initial vaccine series. However, neither the actual duration of humoral protection, nor the clinical factors that impact the strength and duration of this protection are well described.^8–13^ Unlike anti-SARS-CoV-2 antibodies, Spike-specific T-cell responses may be sustained at least up to 6 months after both infection and vaccination.^14^ Poorer concordance between neutralizing antibodies and T cell responses of the adaptive immune system have been associated with severity of disease in individuals ≥ 65 years old.^15^ It remains to be seen whether other clinical factors and co-morbidities negatively impact the strength and duration of the humoral and cellular immune responses after vaccination.

In light of the emerging data on the waning protection of SARS-CoV-2 mRNA vaccines and the clinical benefits of a third dose, the CDC recommends an additional dose after completion of the initial mRNA vaccine series for all adults, and further dosing by age and immunocompromised status.^16,17^ Although advancing age and a personal history of malignancy are associated with decreased immunogenicity to vaccination, those with end-stage renal disease or diabetes mellitus had promising neutralizing antibody responses in other studies.^18–21^ A better understanding of which clinical factors impact post-vaccine immune responses can help guide additional dose requirements over time. We present our results from a longitudinal study on the evaluation of the clinical variables that impact strength and duration of neutralizing antibodies in the sera of individuals vaccinated with Pfizer-BioNTech SARS-CoV-2 mRNA vaccine.

## 2. Methods

### 2.1 Study Design

Starting in December 2020, we conducted a prospective longitudinal study to collect both clinical information and peripheral blood samples from recipients of BNT162b2 (Pfizer-BioNTech) mRNA vaccine, including veterans and healthcare workers at the Veterans Affairs Connecticut Healthcare System (VACHS) located in West Haven, CT, USA. Venous blood was obtained within 48 hours before the first and second doses of vaccine, and then at one month (up to 1.5 months), three months (up to 3.5 months), six months (+/- 2 weeks), and twelve months (+/- 3 weeks) after the second dose. Sera were additionally drawn 1 month after the third (booster) dose (up to 1.5 months), which was on average 10 months after the second dose (SD 0.8). By the 12 month blood draw, only 2 subjects had chosen not to receive their third dose. Venous blood was processed to obtain serum and plasma. which were cryopreserved at -80°C. Clinical and demographic variables were collected via retrospective review of medical records, and included age, race, ethnicity, sex, body mass index (BMI), medical comorbidities (over 80 variables including cardiovascular, pulmonary, oncological, renal, hepatic disease and others), laboratory values (hemoglobin, serum creatinine, hemoglobin A1c (HbA1c), RT-PCR test results for COVID-19 before or during study participation), and concomitant medications. Current steroid use was defined as systemic steroid use for >2 weeks within 1 month before primary vaccine series, or during the study period. Renal function was calculated by estimating glomerular filtration rate (GFR) by using Chronic Kidney Disease Epidemiology Collaboration (CKD-EPI) equation (Table S1).^22^ Subjects whose medical charts were not complete within the VA system because they followed with non-VA physicians, signed release of information for us to obtain their medical records. Per protocol, subjects reported COVID-19 infection, exposure or diagnosis. Three subjects, who had clinically consistent symptoms and exposures but did not get tested, were presumed COVID-positive.

### 2.2 Ethics

The study was approved by the Institutional Review Board at the VACHS. Written informed consent was obtained from each subject.

### 2.3 Evaluation of anti-SARS-CoV-2 neutralizing antibodies

Sera were tested for neutralization activity against SARS-CoV-2 using a single cycle infectivity assay with Spike-pseudotyped virus particles as described previously.^23^ SARS-CoV-2 spike (codon-optimized WA1 or Wuhan-1) pseudotyped lentiviral cores expressing firefly luciferase (FFLUC) were produced. Spike for codon-optimized B1.617.2 (delta variant) were similarly prepared for testing of a small subset of sera (N=36) to compare with WA1-pseudotyped particles at the one-month post third-dose timepoint. These pseudotyped particles were titered on 293T-hACE2 cells seeded in 96 well plates. Each serum sample was tested in duplicate by premixing 75 microliter of four-fold serial dilutions (1:9.21 to 1:603587) with 30 microliter of pseudotyped virus, then adding 95 microliter of this mixture to the 293T-hACE2 cells. After an overnight incubation, 165 microliter of fresh medium was added. After another 48 hours, cells were lysed and luciferase luminometry performed. Curve-fitting using GraphPad PRISM was employed to calculate the neutralization titer half-maximal inhibitory concentration (IC50 value) for each sample at each timepoint.

### 2.4 Statistical analysis

#### Variable Selection

Subject morbidity data were grouped based on organ system involved, and clinical characteristics (Table S1). Undetectable IC50 values (<9.21 μg/mL) were changed to 9.21 for statistical analyses. The IC50 values were then log-normalized. Paired t-test analysis was used to compare log-normalized IC50 values at various timepoints: one, three, six, ten, and twelve months with baseline log_2_IC50 values before vaccination. To compare IC50 values in subjects with prior COVID-19 at 12 months, an unpaired t-test was performed after log normalization.

To assess the neutralization assay over time and as a comparison with baseline values, the fold-change (Figure 1B) was calculated by dividing the IC50 at each timepoint by the baseline IC50. As shown in the y-axis of Figure 1B, the fold changes were found to follow a logarithmic distribution and were thus log-normalized. These steps to calculate log_2_fold-change (LFC) are shown in the equation below and were used in all univariate and multivariate analyses (Figures 2-4).

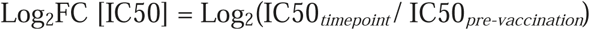

**Figure 1.**
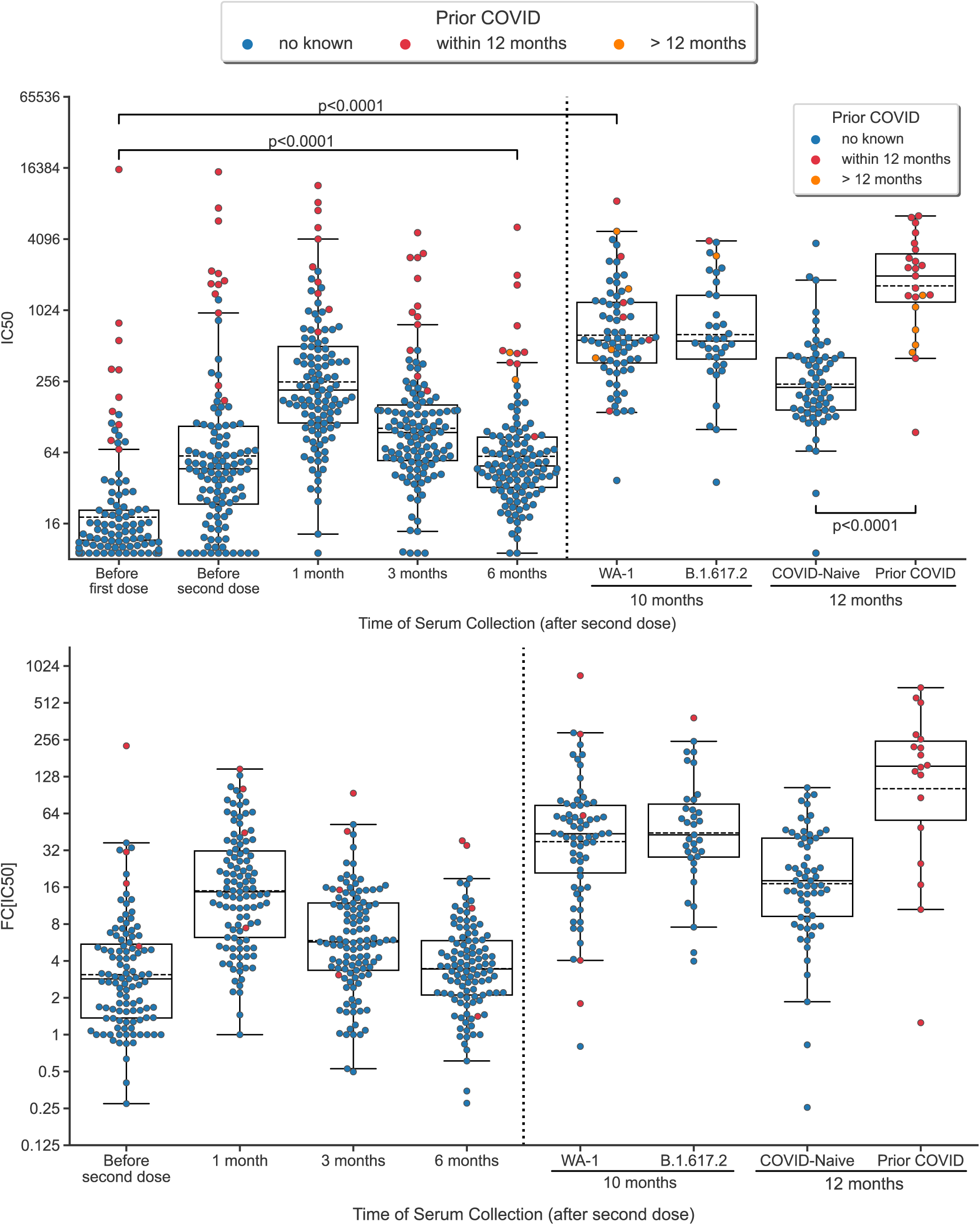
Neutralization antibody titers following vaccination. (1A) Swarmplot/boxplot of IC50 plotted over time before and after vaccination, second dose, and third dose. Boxplot shows median, lower/upper quartile, and extremes. (1B) Swarmplot/boxplot of fold-change (FC[IC50]) over pre-vaccination IC50 plotted over time after the first dose of vaccine. P-values are derived from paired t-test. Outliers, values further than 1.5 times the interquartile range (Q_1_ – 1.5*IQR and Q_3_ + 1.5*IQR), are shown beyond the boxplot. Subjects with COVID prior to first dose of vaccination were excluded in figure 1B because it artifactually lowers the fold-change.The 10 month time point was 1 month post third-dose. The 12 month timepont was 3 months post third-dose.

Log_2_ΔIC50 was also considered as an alternate outcome variable in which the pre-vaccination IC50 was subtracted from the IC50 at each timepoint. Ultimately, LFC was chosen to adequetely normalize the outcome for parametric statistical tests and consistent reporting with prior studies. However, using LFC results in a measurement artifact in which higher pre-vaccination IC50 values are associated with lower increases in LFC.^24^

In our analysis, we used the highest LFC as a measure of ‘strength’ of the humoral response. Additionally, we calculated the LFC of titers at six and twelve months compared with titers at pre-vaccination to represent the ‘duration’ of the response after the primary series and third dose respectively.^24^ Positive LFC values represent increase in IC50 compared with pre-vaccination IC50 and negative values represent a decrease in IC50.

#### Univariate analysis

Continuous variables (Xi) were used in analyses with outcomes log2(FC) (neutralization titers). Pearson or Spearman correlation tests were utilized depending on the distribution of the response variable:

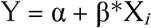

In this equation, Y represents log_2_FC. R values reported represent the strength and direction of correlation.

For dichotomous categorical variables, if both subgroups maintained normal distributions (p>0.10 using Shapiro-Wilk test) of the outcome variable, then we reported the results of two-sided student’s (independent) on t-test (one-sided). If, however, at least one subgroup did not maintain a normal distribution (p<0.10), then we used the non-parametric test, Wilcoxon rank-sum test (one-sided). All univariate analyses were conducted using the Python (v3.8.5) statsmodels package.^25^

#### Multiple regression analysis

Variables (X_i_) were added and subtracted in a stepwise manner to optimize the Akaike information criterion, an established metric quantifying the trade-off between underfitting and overfitting the data.^26,27^

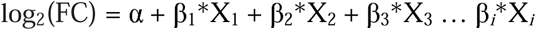

To account for the artifactual negative correlation between pre-vaccination IC50 and log_2_FC values, log_2_pre-vaccination IC50 values was also added as a variable in our analysis. Reported are the coefficient estimates representing the slope of each variable, βi.Non-signficant variables (p>0.10) were excluded from the figure. Positive and negative values (full range between negative infinity and positive infinity) represent positive and negative association between clinical variable and response. The intercept represents y-intercept of the model (i.e., younger subjects without co-morbidities and higher GFR response variables). Goodness of fit was analyzed using Q-Q plots. Multiple regression analysis was conducted using R (v4.1.1) with the MASS package.

### 2.5 Visualizations

All univariate association figures were created using the Python package seaborn. Forest plots, illustrating multiple linear regression models were generated using R package forest plot.

## 3. Results

We included 124 subjects (91 veterans, 33 healthcare workers), of whom 74% were male, 66% were white, 93% non-hispanic, and 60% were over the age of 65 years (Table 1). Ten subjects had COVID-19 diagnosed by RT-PCR prior to the first dose of vaccine and were thus excluded from the univariate and multivariate analyses. Two subjects developed COVID-19 before month 6 after the second dose of vaccine, and their 6-month IC50 values were excluded from analyses. All 124 subjects had sera serially collected until 6 months after their second dose. Of these, 72 subjects had sera drawn 1 month after their third dose (10 months post second-dose), and 88 subjects had sera drawn 3 months after third dose (12 months post second-dose) by the time of this publication. Two individuals did not receive their third dose by 12 months, one of whom contracted COVID. By 12 months, 23 subjects had contracted COVID-19 and COVID-19 was added as a variable into the multivariate regression.

**Table 1.** Cohort Demographics and Morbidities (n=124)

|  | n (%) |
| --- | --- |
| Male | 91 (73.4) |
| Age, in years |  |
| 20-64 | 50 (40.3) |
| 65-95 | 74 (59.7) |
| Race |  |
| White | 82 (66.1) |
| Black | 22 (17.7) |
| Other | 20 (16.1) |
| Ethnicity |  |
| Non-Hispanic | 108 (87.1) |
| Hispanic | 8 (6.5) |
| COVID-positive |  |
| No | 110 (88.7) |
| Yes, before first dose | 10 / 121 (8.3) |
| Yes, at 1-month serum collection | 10 / 121 (8.3) |
| Yes, at 3-month serum collection | 12 / 121 (9.9) |
| Yes, at 6-month serum collection | 12 / 121 (9.9) |
| Yes, at 10-month serum collection | 10 / 72 (5.8) |
| Yes, at 12-month serum collection | 23 / 88 (26.1) |
| Chronic Heart Disease | 77 (62.1) |
| Lung Disease | 67 (54.0) |
| Current Steroid Use | 19 (15.3) |
| Liver Disease | 8 (6.5) |
| GFR in mL/min/1.73m<sup>2</sup>, mean (SD) [1] | 69.6 (31.2) |
| >60 | 87 (70.2) |
| 30-60 | 18 (14.5) |
| <30 | 17 (13.7) |
| Malignancy |  |
| Solid | 31 (25) |
| Hematological | 5 (4) |
| Both | 2 (1.6) |
| Diabetes Mellitus | 39 (31.5) |
| TIA-CVA | 13 (10.5) |
| Dementia | 5 (4.0) |
| HIV-Transplant-Rheum | 18 (14.6) |
| HbA1c, median [Q1,Q3] [2,3] | 5.6 [5.2,6.1] |
| BMI, median [Q1,Q3] [2,3] | 28.4 [24.8,31.5] |

[1] Multimodal distributions (Hartigan's Dip Test)
[2] Non-normal distributions (Shapiro-Wilk Test)
[3] Far outliers in Tukey test

### 3.1 Time since vaccination and anti-SARS-CoV-2 neutralizing antibody titers

Among the timepoints tested, IC50 values were highest at one month after the second dose of vaccine, with a median fold change (FC) of 14.1 compared to pre-vaccination titers, thereafter declining to a median FC of 5.6 and 3.3 at three months and six months, respectively. Consistent with previous reports^24^, we found that higher pre-vaccination IC50 values were associated with a lower FC response at one month (R=-0.27, p=0.005) and six month timepoints (R=-0.63, p<0.0001) (Figure 2). Furthermore, we found that individuals with stronger response strength (higher FC at one month) also had maintained duration of vaccination response (higher FC at six months) (R=0.72, p<0.0001).

**Figure 2.**
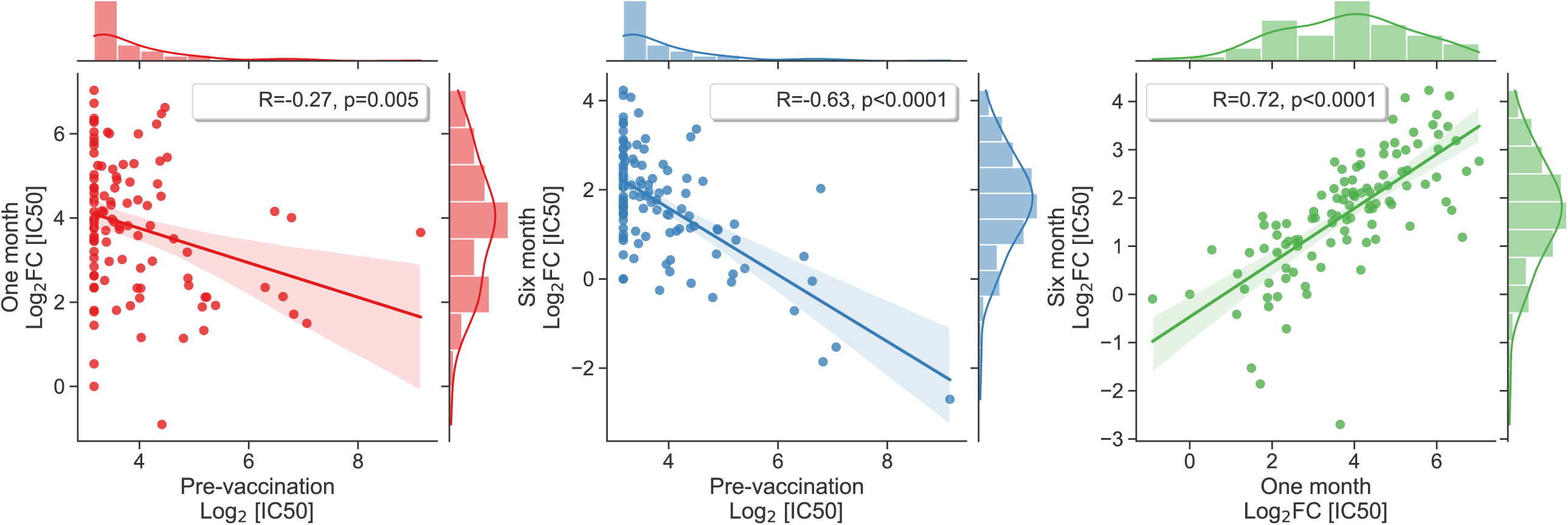
Association between pre-vaccination IC50 and fold-changes in neutralizing antibody response. Scatterplot and Pearson correlation coefficients between pre-vaccination IC50 and fold-change response at one-month and six-months. Not shown: similar correlation with pre-vaccination IC50 found at 10- and 12-months, (R=-0.40, p=0.0015 and R=-0.54, p<0.0001 respectively).

At 1 month post third-dose (10 months post second dose), the median FC increased to 41.5. (Figure 1). This peak was significantly higher than the peak response measured 1 month post second-dose (p<0.0001). Although the median FC for WA-1 and B1.617.2 at 1 month post third dose was 41.5 and 41.2 respectively, the FC for WA-1 was consistently higher for each subject compared to B1.617.2 in a paired analysis of 36 subjects (p=<0.0001). Pre-vaccination IC50 maintained a negative correlation with fold-change at 1 month (R=-0.40, p=0.0015) and 3-months (R=-0.54, p<0.0001) post third-dose. FC response at six months positively correlated with 1-month FC (R=0.35, p=0.0061) and 3-month FC (R=0.50,p<0.0001) post third-dose response.

### 3.2 Hybrid immune response

By the 12-month timepoint (3 months post third-dose), 23 subjects had contracted COVID-19. The IC50 was significantly higher in these subjects (median FC 132) compared to COVID-19 naïve (median FC 18.1; p<0.0001). Those with COVID-19 >12 months prior, had signicantly lower IC50 values than those with COVID-19 within 12-months (p=0.004), but still significantly higher than the COVID-naïve (p=0.001).

### 3.3 Correlation of neutralizing antibody response to clinical variables

#### Univariate analysis

To understand the role of demographic and clinical factors in the strength and duration of the humoral response, univariate analysis was conducted after excluding subjects with prior COVID-19 diagnoses. Results are shown in Figures 3 and S1.

**Figure 3.**
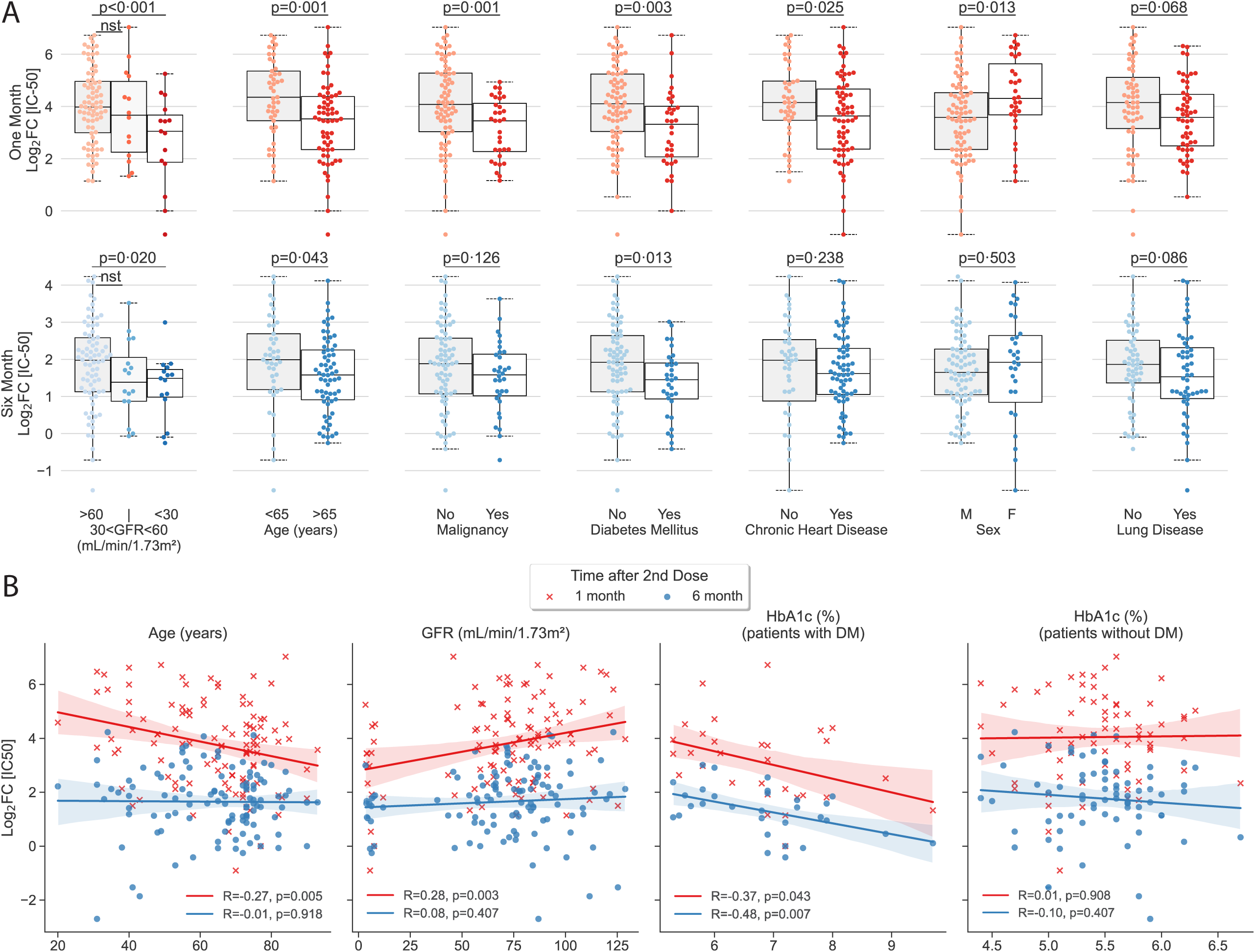
Univariate analysis showing clinical factors that correlated significantly with neutralizing antibody titer peak and duration at 6 months. (3A) Categorical analysis between significant (p<0.05) and trending variables (p<0.10) of vaccination response at one month (left) and six months following second dose of the vaccine. (3B) Scatterplot illustrating continuous variables: age, glomerular filtration rate, and hemoglobin A1c plotted against vaccination response at one month (red crosses) and six months (blue circles). Correlation analyses of hemoglobin A1c was conducted separately for patients with and without diabetes. Colored lines represent lines of best fit, with shading showing 95% confidence intervals. Subjects with prior COVID-19 diagnosis were excluded from univariate analysis. NS represents non-significant or trending association.

#### Multiple Linear Regression

To account for the multi-morbidity of our subjects, we conducted a stepwise multiple linear regression both for the response strength and response duration (Figure 4). BMI was intentionally excluded due to collinearity with chronic heart disease. Because of the strong negative correlation between pre-vaccination IC50 values and response measured as log_2_FC, the baseline log_2_IC50 value was added to the stepwise regression. Unsurprisingly, higher pre-vaccination IC50 maintained a significant association with lower fold-change values at one, six, ten, and twelve months consistent with our univariate analyses, as well as, previously reported neutralization assays.^24^

**Figure 4.**
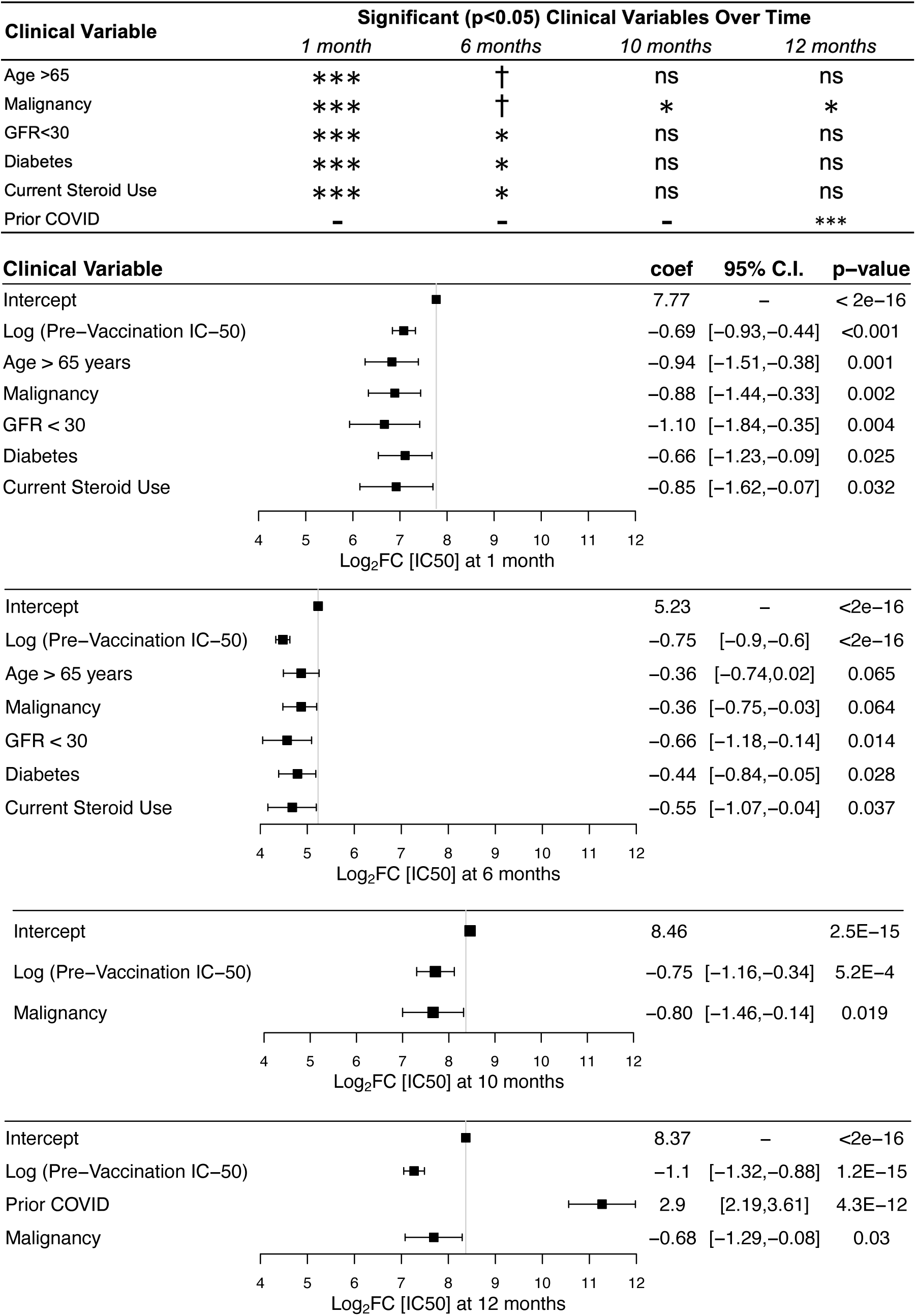
Multiple linear regression of clinical variables associated with neutralizing antibody response. Summary plot (top) showing variables retained in the stepwise regression with corresponding significant p-values. Non-significance denoted by ‘ns’, p-value between 0.10 and 0.05 denoted by †, p-value between 0.05 and 0.01 denoted by *, p-value between 0.01 and 0.01 and 0.001 denoted by **, p-value less than 0.001 denoted by ***. Log_2_FC = log normalized fold-change over pre-vaccination IC50. Forest plots showing multiple linear regression variables of response at one month, six months, ten months, and twelve months following administration of second dose. Variables not reaching significance not shown in the figure are sex, race, chronic heart disease, lung disease, liver disease, current steroid use, cerebrovascular disease, HIV-Transplant-Rheum, and dementia. Intercept represents baseline comparison values: age less than 65 years, GFR greater than 60 mL/min/1.73m^2^, no steroid use, and no comorbidities. The 10 month time point was 1 month post third-dose. The 12 month timepont was 3 months post third-dose.

Significant clinical factors associated with decreased strength of response at one month after primary vaccine series completion, included age >65 years (β=-0.94, p=0.001), malignancy (β=- 0.88, p=0.002), GFR < 30 mL/min/1.73m^2^ (β=-1.10, p=0.004), current steroid use (β=-0.066, p=0.032), and diabetes mellitus (β=-0.57, p=0.032). (Figure 4). GFR < 30 mL/min/1.73m^2^ (β=- 0.66, p=0.014), current steroid use (β=-0.55, p=0.037), and diabetes mellitus (β=-0.44, p=0.028) were significantly associated with decreased duration of response at 6-months. Meanwhile, age >65 years (β=-0.36, p=0.065) and malignancy (β=-0.36, p=0.064) showed a trend towards reduced duration without meeting significance (Figure 4). Malignancy significantly decreased the post third-dose response at one month (β=-0.80, p=0.019) and three months (β=-0.68, p=0.03).

## 4. Discussion

SARS-CoV-2 vaccines have played a key role in curbing the spread, morbidity, and mortality from COVID-19. With rising concerns regarding the duration of protection after vaccination, efforts to understand the optimal timing and number of additional doses of vaccine are underway. It is imperative to understand clinical factors that impact the strength and duration of the immune responses to these vaccines, and the role of adaptive immune responses on duration of protection. Previous studies have examined one or two clinical factors to assess the effect of vaccination on immune response.^18–21^ In this study, with full access to medical records, we evaluated multiple clinical factors with multivariable regression in order to identify which clinical variables had the most significant negative (or positive) impact on the strength and duration of immune response over the course of 1 year.

This study was conducted on a U.S.-based population of veterans and healthcare workers, which offers a unique study demographic. About half of our sample population for this cohort was >65 years old and the majority was male with multiple co-morbidities. The predominantly older male population typical of veterans is also a predisposed demographic group to suffer severe/fatal COVID-19, especially if there are coexistent co-morbid conditions.^28, 29^ It is critical to determine how protective and persistent the vaccine response will be in individuals at high risk for contracting severe COVID-19. We included healthcare workers in this analysis to enhance the diversity of the study population in terms of demographic and clinical factors.

In this cohort, we found that among the timepoints tested, IC50 values peaked at one-month after completion of the primary series of BNT162b2 mRNA vaccine, with the six-month median level at one-fourth of the peak level. Our data suggest that age and malignancy play a role in reducing the initial peak response, but do not significantly impact the duration of response at 6-months, although they trend towards significance.

The third dose of mRNA vaccine has been shown to boost the waning immune response. Our results confirm that the immediate post third-dose response was robust against variants WA-1 and B1.617.2, and other studies have demonstrated protection against the Omicron variant as well.^30,31^ At one and three months after the third dose, a history of malignancy was the only clinical variable that significantly reduced the IC50. Given that the level of neutralizing antibody titers have been shown to be highly correlated with immune protection,^7^ these findings support the use of booster doses, with prioritization for specific, vulnerable populations, in particular those with a history of malignancy, as other studies suggest.^32,33^

We found that SARS-CoV-2 infection enhanced the neutralization antibody response to third dose, and that this effect was most prominent if the subject had COVID-19 within 12-months, but both the COVID-19 naïve and those with prior COVID-19 infection significantly benefited from the additional dose. This finding is supported by prior literature highlighting the synergistic response between the immune response from vaccination and infection with various variants, facilitated by B- and T-cell activation.^34,35^

Antibody titers after a third dose of mRNA vaccine have been shown to correlate with pre-existing B-cell frequency and may benefit from T- and B-cell memory that allows for efficient, rapid, and high-level production of neutralizing antibodies.^36^ The short-term effectiveness of a 4^th^ dose at preventing COVID-19 infection, but the potential longer protective effect against severe disease, have been suggested to support that certain populations may benefit from additional immune memory priming.^37^ Further study will be needed to understand how many doses are beneficial over time, whether immune memory will offer sufficient protection, and whether different clinical factors will impact that immune memory.

This study has some important limitations. Our sample size is relatively small, and limited in ethnicity distribution. More generalizable results could be generated from larger distributions of ethnicity. Subjects who had 12-month IC50 analysis had received third dose three months before. While this provided an insight into clinical variables impacting duration of third dose response, evaluation of 12-month IC50 without third dose would enrich the information further. Similarly, we defined strength of response as the highest IC50 obtained by each individual. Since the timepoints of collection were predetermined, it is possible that the true peak response occurs before or after the 1-month timepoint. While this may affect fold-change values, it will likely not alter the correlation of strength of response with clinical factors.

In conclusion, while age >65 years negatively impacts the initial strength of the humoral immune response as quantified by levels of neutralizing antibodies, clinical comorbidities of end-stage renal disease, diabetes mellitus, and steroid use negatively impact the initial strength and duration of the humoral immune response against SARS-CoV-2 at 6 months following the primary series. The response to the third dose was universally robust suggesting a sustained immune memory response, except in those with malignancy, suggesting that the latter population may benefit from further doses of vaccine. Prior COVID-19 enhanced vaccine-generated response, but the effect wanes with time. Further exploration is needed into the durability of immunologic memory of SARS-CoV-2-specific T- and B-cells for vulnerable populations.

## Data Availability

All data produced in the present work are contained in the manuscript.

## Acknowledgements

We thank the Veterans and healthcare workers who have donated their samples to make this study possible.

