## Supplementary material for "Clinical Variables Correlate with Serum Neutralizing Antibody Titers after COVID-19 mRNA Vaccination in an Adult, US-based Population": Table S1

| **Table S1. Categorization criteria for morbidities in our cohort** | |
| --- | --- |
| **Clinical Variable** | **Morbidity Criteria** |
| Chronic Heart Disease | Coronary artery disease, arrhythmias including atrial fibrillation, hypertension, congestive heart failure, peripheral vascular disease, structural heart disease |
| Lung Disease | Chronic obstructive pulmonary disease, asthma, interstitial lung disease, obstructive sleep apnea, (malignancy not included) |
| Current Steroid Use | Systemic steroid use for >2 weeks in the following time period: within one month before the first dose of vaccine through the study period. |
| Liver Disease | Alcoholic hepatitis, untreated viral hepatitis, nonalcoholic steatohepatitis, hemochromatosis, cirrhosis (decompensated and compensated). |
| Cerebrovascular Disease | Stroke (including history of transient ischemic attack) |
| Cancer | Solid organ or hematologic |
| HIV-Transplant-Rheum | HIV, Solid or stem-cell transplant recipient on active immunosuppressive medication, Rheumatological or autoimmune diseases with known compromise of immune competence |
| BMI | Body mass index, in kg/m^2^, within one month before first dose of vaccine |
| HbA1c | Highest hemoglobin A1c (%) within the study period. |
| Renal Function | Baseline GFR during the study period, in mL/min/1.73m^2^, as calculated by CKD-EPI formula |
