## Supplementary figures and images for "Clinical Variables Correlate with Serum Neutralizing Antibody Titers after COVID-19 mRNA Vaccination in an Adult, US-based Population"

### Figure S1

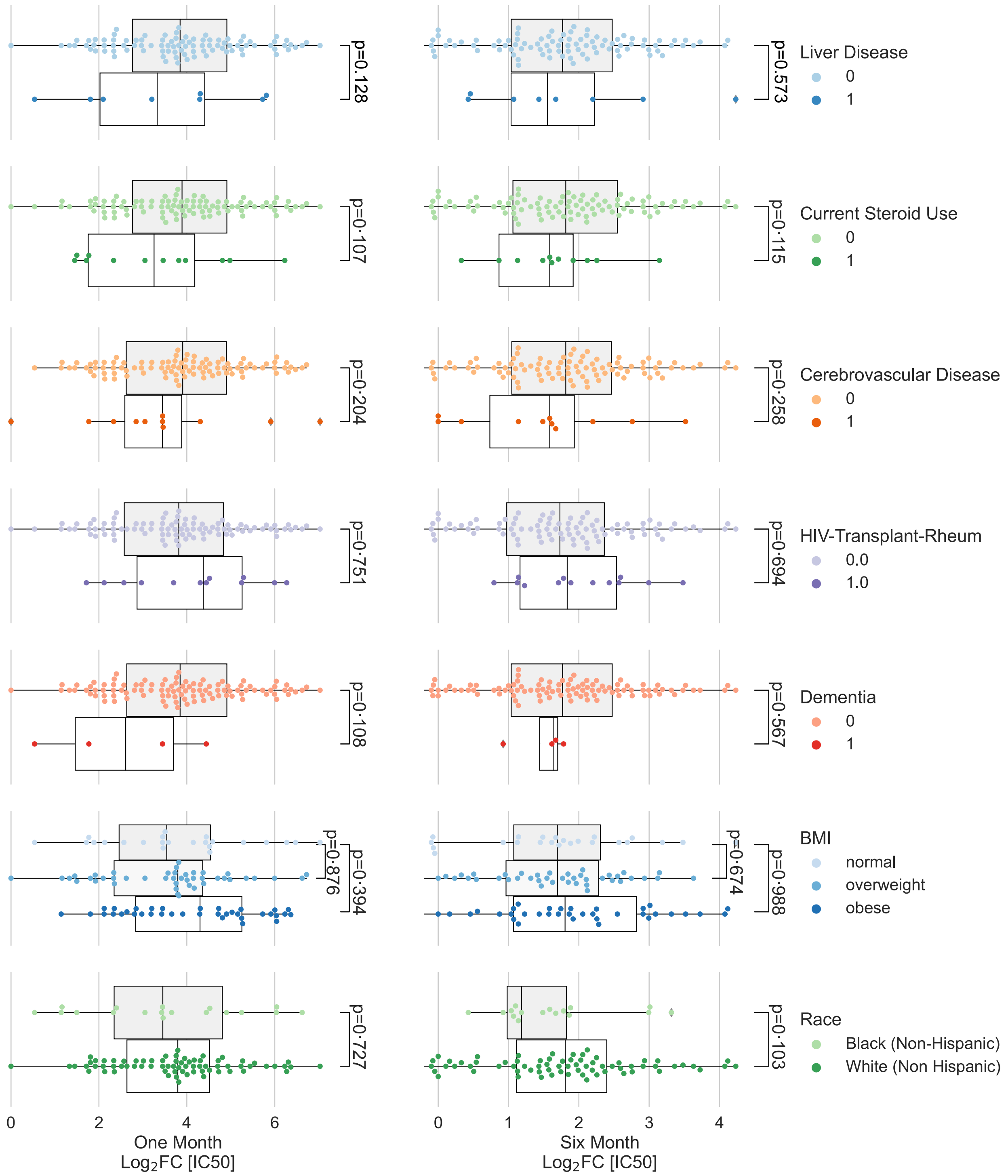

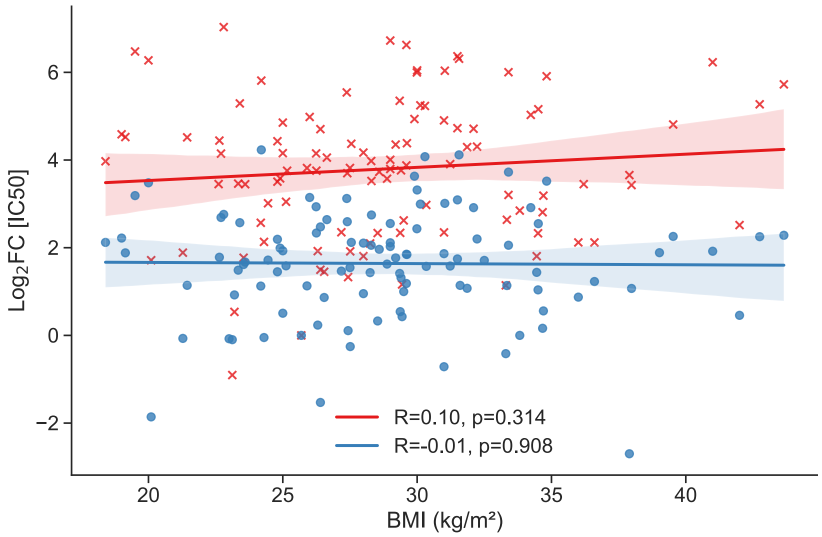
